# Study protocol for an inception cohort investigating possible predictors of systemic steroids effectiveness in treatment of acute sciatic pain

**DOI:** 10.1101/2021.02.24.21252392

**Authors:** Guilherme Henrique Porceban, Renato Salvioni Ueta, João Carlos Belloti, Cláudio Antônio da Costa Neto, Alexandre França Filho, Fabio Antônio Vieira, Eduardo Barros Puertas, Marcel Jun Tamaoki

**Author notes:** Corresponding author: Guilherme Henrique Porceban, Departamento de Ortopedia e Traumatologia, Rua Napoleão de Barros 715, Vila Clementino, 04024-002 São Paulo – SP, Brazil. /.

## Abstract

**Introduction:** Sciatic pain secondary to nerve root compression occurs in approximately 1% of the general population in the United States, which represents enormous costs related to the treatment and loss of function of symptomatic individuals.

Acute radicular pain is predominantly caused by herniation of the intervertebral disc but can also be caused by degenerative changes. Compressive and inflammatory mechanical factors are interrelated in the pathophysiology of symptoms.

Although radiculopathy is self-limited in most cases, in its acute stage it is associated with painful symptoms and loss of essential function. The first-line treatment usually employed is a conservative approach, including a short rest period and use of common analgesics and nonsteroidal anti-inflammatory drugs.

As an alternative to the conservative approach, the systemic administration of oral corticosteroids, such as prednisone, are widely used. However, the current literature shows contradictory results for this treatment in terms of improvement of pain and function.

The present study hypothesizes that treatment with prednisone is effective in the treatment of acute sciatic pain in patients with social, clinical, and demographic characteristics favorable to this treatment.

**Objectives:** the main objective of the present study is to identify predictors, both clinical and imaging, of a positive response, in terms of both function and pain intensity, to prednisone treatment in patients with acute sciatic pain.

**Methods and analysis:** the present study will include a cohort of patients, with a diagnosis of acute sciatica, who will receive treatment with oral prednisone. The pain and functional scores before and after treatment will be compared. Thereafter, the social, clinical, and radiographic characteristics of responsive patients will be compared to those of patients who did not respond well to the treatment.

**Ethics and dissemination:** the study received the approval of Federal University of São Paulo Research Committee (4.232.193) and Research Ethics National Committee (CONEP).

**ARTICLE SUMMARY:** Strengths and limitations of this study

- The study is based on samples from a large population recruited from a reference facility for the treatment of acute sciatic pain.
- There is potential for optimization of treatment response with oral corticosteroids.
- The study will be conducted with a population with defined clinical characteristics.
- Due to the relatively short follow-up period, the project will not evaluate alternative treatments, such as surgery.

## INTRODUCTION

Low back pain is an important cause of absence from professional activities and is the sixth greatest cause of loss of quality of life and function worldwide among diseases; in some regions, it tops the list.^1^ It is estimated that low back pain is responsible for 139 million medical appointments per year in the United States.^2^

In approximately 12% of individuals who present with back pain,^3^ there is an association with radiculopathy, which corresponds to the pain that follows the dermatome or myotome of the affected nerve root (sciatica pain), associated with signs of increased tension in the nerve root. This can be identified through clinical examination maneuvers. In the general population, the prevalence of sciatic pain is estimated to be around 1%.^4^

The most prevalent cause of acute radiculopathy is lumbar disk herniation.^5^ Nucleus pulposus extrusion due to lesions of the annulus fibrosus leads to mechanical compression and triggers local inflammatory responses, mediated by proteins such as alpha tumor necrosis factors (TNFα) and phospholipase A2, which attract macrophages and lymphocytes to the compressed region and increase local inflammatory processes, causing pain and, in certain situations, paresthesia and motor deficits.^6–9^ The exact contribution of mechanical and inflammatory causes in acute sciatic pain is controversial.

Traditional treatment for acute radiculopathy involves resting for a short period and use of nonsteroidal anti-inflammatory drugs (NSAIDs), muscle relaxants, and opioids.^10–12^ In about 50% of patients, acute sciatica regresses within 6 weeks, and 90% of the patients recover within 12 weeks.^13^ Although in the vast majority of patients sciatica is a self-limiting disorder, these individuals are mostly professionally active, and the financial losses caused by absence from their professional activities are enormous. An alternative for the conventional treatment is the lumbar epidural steroid injections (LESI), an interventional procedure for patients whose initial treatment has failed or those who presents severe sciatica pain or disfunction. The number of interventions for lumbar radicular pain has increased since 1990 and nowadays they constitute a widely used treatment, although there is a growing body of reports describing severe adverse effects of such procedure.^12-13^

Systemic administration of oral corticosteroids is recommended by the guidelines of the North American Spine Society and the American Academy of Orthopedic Surgeons.^14^ This treatment can be conducted by general physicians in low complexity health services such as primary and secondary unities.

The mechanism of action of steroids for pain relief has not yet been fully determined. It is believed that steroids act as neural blockers by altering or interrupting nociceptive stimuli and the reflex mechanisms of the afferent fibers.^15,16^ Furthermore, corticosteroids have been proven to reduce the inflammatory response by inhibiting both the synthesis and release of proinflammatory mediators.^17–19^

Several studies have sought to evaluate the effectiveness of oral corticosteroids in relieving acute sciatic pain, but its effectiveness has not been fully demonstrated and remains controversial in terms of evidence. In the most relevant and impactful of these studies,^20^ a group of patients with acute sciatic pain who underwent a short-term course of treatment with prednisone showed an modest improvement in function at 3 weeks and 52 weeks after the treatment when compared to the placebo group.

Studies with varying methodologies and sample sizes have reported different results on the effectiveness of this class of drugs. A systematic review involving smaller studies with different treatment methodologies and protocols^21^ concluded that the effectiveness of corticosteroids is not superior to that of placebo for acute sciatica pain, and it also has more side effects. This discrepancy in the literature is also found on the subject of transforaminal corticosteroid injections.^22–24.^ However, systematic reviews, meta-analyses, and cohort studies were able to identify some clinical and radiological characteristics of patients who benefit from these treatments.^25,26^ This eventually lead to the use of this therapeutic modality for the group of patients who present those parameters.

Despite the controverse evidence in the literature on the effectiveness of corticosteroids, it is estimated that approximately 5% of patients with acute sciatic pain use them^27,28^; their use is particularly popular among patients with severe pain intensity and in places where the health system is unable to support more complex and invasive procedures.

Currently, no study published in the literature has attempted to correlate the sociodemographic, clinical, and radiological characteristics of patients with acute radiculopathy to the effectiveness of oral systemic corticosteroids in treating this condition.

By acting on the inflammatory component of acute radicular pain, corticosteroid therapy may have more benefits for patients whose pain is mainly related to this mechanism. For example, those that present parameters such as a short period since the onset of pain, images that do not show marked foraminal stenosis, and low intensity of associated axial pain.

Identifying the characteristics that can predict a favorable response to systemic corticosteroids can facilitate the selection of patients who would potentially benefit from the use of this class of drugs, while preventing those with low probability of improvement in pain and function from being exposed to these drugs’ side effects.

Currently, no study published in the literature has attempted to correlate the sociodemographic, clinical and radiological characteristics of pacientes with acute sciatica pain to the effectiveness of oral systemic steroids in treating this condition.

### Objectives

The objective of this study is to evaluate clinical, demographic, and imaging characteristics as possible predictors of the success, in terms of function and pain, of oral corticosteroid treatment in patients with acute radiculopathy.

A secondary objective of the study is to perform a cross-sectional epidemiological assessment of individuals seeking emergency care for acute sciatic pain in a regional reference. The intensity of pain and loss of function of these patients at the first visit will be evaluated and compared with those reported in studies on populations of different countries. To date, there is no epidemiological studies involving the Brazilian population in the current literature.

## METHODS AND ANALYSIS

### Study design and patient involvement

This is a prospective cohort study of patients with acute radicular pain undergoing a course of systemic corticosteroid therapy.

Patients will be recruited from the Orthopedic and Spine outpatient clinic of the São Paulo Hospital (SPH) of the Federal University of São Paulo, Brazil, and from an affiliated hospital, the Barueri Municipal Hospital (BMH), located in a nearby city. Patients with a clinical presentation compatible with acute radicular pain and who meet the eligibility criteria, described in Table 1, will be included in the study.

**Table 1.** Inclusion and Exclusion Criteria

| Table 1 - Inclusion and Exclusion Criteria |  |
| --- | --- |
| Inclusion | • Age > 18 years<br>• Clinical history compatible with severe acute (less than 3 months from onset) sciatica pain (ODI>40) |
| Exclusion | • Patients who refuse to participate in this study<br>• Patients who refuse to take medications, lost follow-up or has contra-indications for MRI<br>• Co-morbidities that contra-indicates the steroids use<br>• Clinical history of active infections, tumors, spine trauma, previous spine surgery or pain intervention less than 3 months ago<br>• Clinical history compatible with cauda equina syndrome |

The patients will receive an informed consent form (ICF), and if they agree to participate, they will complete questionnaires for quantitative/qualitative assessment of pain and function, namely the Visual Analogue Scale (VAS) and the Oswestry Disability Index (ODI).^29–31^ They will also undergo magnetic resonance imaging (MRI).

At this stage, patients will receive material for basic education, in language appropriate to laypeople, about their disease and how the study can help to optimize treatment. Still, they will be encouraged to contribute to the study, when reporting which outcomes are considered most relevant for them to consider the treatment as effective.

The research team will review the information obtained through these efforts in order to consider as priority those most frequently reported by the participants, be it, for example, the quick resumption of the work routine, pain relief, resumption of sport, among others.

Initially, based on similar studies available in the literature, ^15-19^ the authors assumed as primary outcome of the study is improved pain and functional status, represented as VAS and ODI scores, respectively, three weeks after the end of corticosteroid treatment.

The patients will be considered to have responded to treatment if they achieve a 50% reduction in VAS score or a 7.1% reduction in the ODI score; these parameters were set previous studies that considered these values as minimum indicators of improvement in quality of life.^32,33^

Thereafter, they will be divided into two groups, according to the success or failure of the treatment. These groups will be compared in terms of the social, clinical, and radiological parameters described in the collected data section.

### Collected data

#### Demographic data

At the time of a patient’s first consultation, their demographic data will be collected for subsequent comparison, in order to evaluate possible predictors. These data include the following:

a. Age
b. Gender
c. Ethnicity: white, mixed, black, or Asian
d. Body Mass Index (BMI)
e. Professional activity: active or retired/unemployed
f. Smoking: yes/no
g. Secondary earnings

#### Clinical data

Patients eligible to participate in this study will undergo a clinical evaluation and physical examination conducted by the main author of this study or by one of the co-authors. The parameters to be obtained from this evaluation include:

a. Time of onset of symptoms: < 1 week, 1 week to 1 month, and between 1 and 3 months
b. Similar previous episode of pain
c. ODI score according to the questionnaire version validated for the Portuguese language^29^
d. VAS score for low back pain and radicular pain^30^
e. Side of the body affected
f. Presence of motor deficit, defined as muscle strength ≤ 3 in the myotome corresponding to the spinal root, according to the Medical Research Council scale for the evaluation of muscle strength,^38^ assessed by physical examination
g. Presence of tactile sensitivity deficit, defined as a reduction in the sensitivity of the affected dermatome compared to the contralateral side, assessed by physical examination

#### Magnetic Resonance Imaging assessment

T1 and T2 weighted MRI will be performed, and images will be accessed via a picture archiving and communication system. These images will be evaluated by two experienced orthopedists, specializing in spine surgery, and a radiologist, specializing in the musculoskeletal system; all evaluators will be blinded to the patient’s clinical picture and response to prednisone treatment.

The analysis of MRI in patients with lumbar disk herniation will be conducted similar to previous studies about sciatica pain.^34^ Degenerative alterations, including osteophytes in the terminal spinal plate, facet hypertrophy, and thickening of the ligamentum flavum, that probably contribute to foraminal stenosis will be described.

In patients with a herniated disk, the morphology of the hernia will be evaluated in axial and sagittal slices. Initially, hernias in axial slices will be classified according to their position into central, in the lateral recess, or foraminal. Additionally, from the axial slices, the horizontal and vertical diameters of the hernia and the area of the spinal canal will be calculated. The ratio between the vertical diameters of the hernia and spinal canal will be estimated as a magnitude of the obliteration of the canal by the herniated disk.

The degree of foraminal stenosis and consequent root compression will be assessed from the T1-weighted sagittal slices based on the classification proposed by Lee et al.^35^

In short, the imaging parameters evaluated will be as follows:

a. Predominant compression site: anterior, posterior, or mixed
b. Degenerative alterations (osteophytes, facet hypertrophy, thickening of the ligamentum flavum): present or absent
c. Herniation of the nucleus pulposus: present or absent
d. Vertical hernia/spinal canal ratio - a relationship between the longest vertical length of hernia and the longest vertical length of the spinal canal, in millimeters (Figure 1)
e. Horizontal hernia/spinal canal ratio - a relationship between the longest horizontal length of hernia and the longest horizontal length of the spinal canal, in millimeters (Figure 2)
f. Degree of foraminal stenosis according to classification by Lee et al.^35^

**Figure 1.**
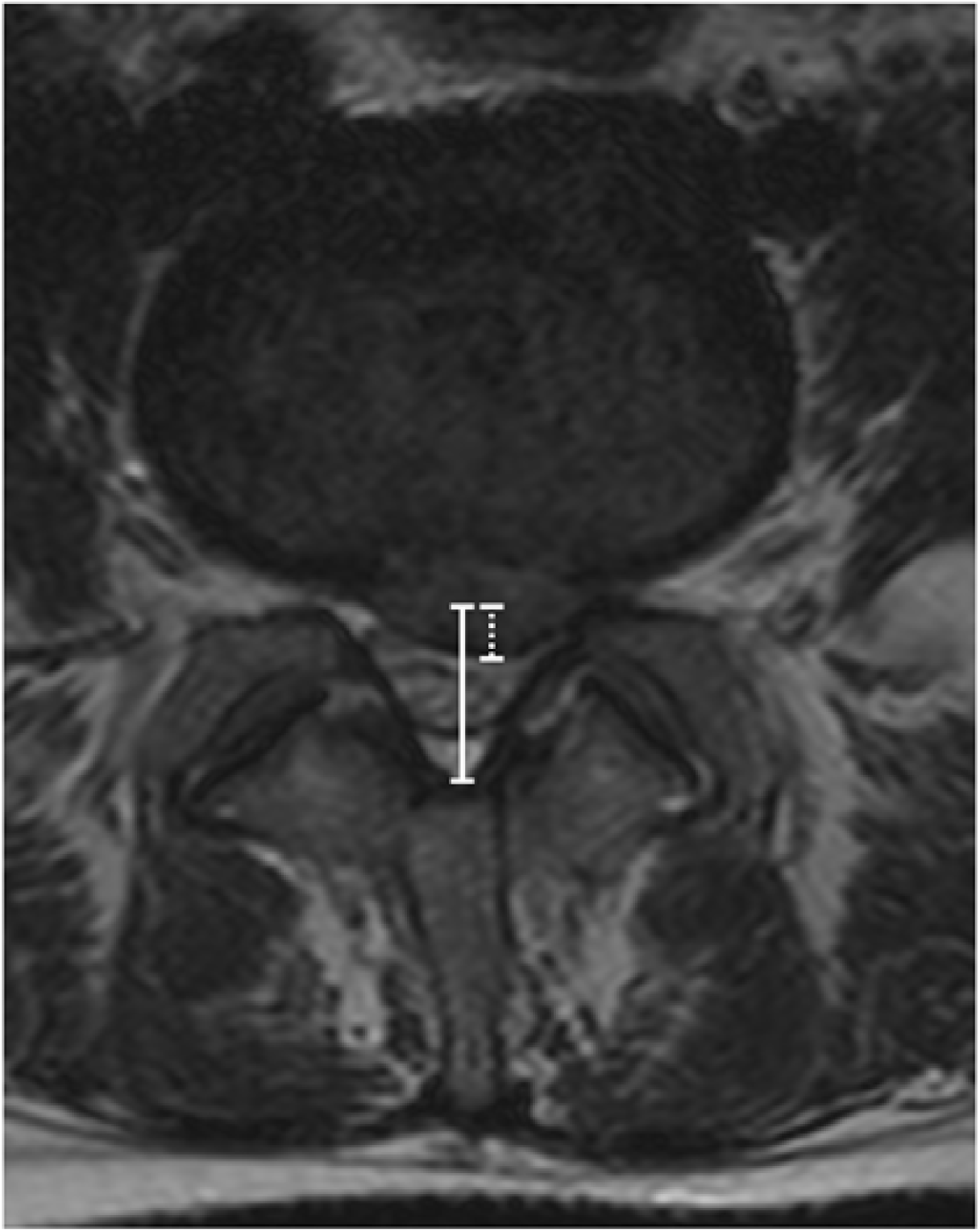
Measurements of the vertical diameter of the disk herniation (dashed line) and the vertical diameter of the spinal canal (continuous line) in the axial plane.

**Figure 2.**
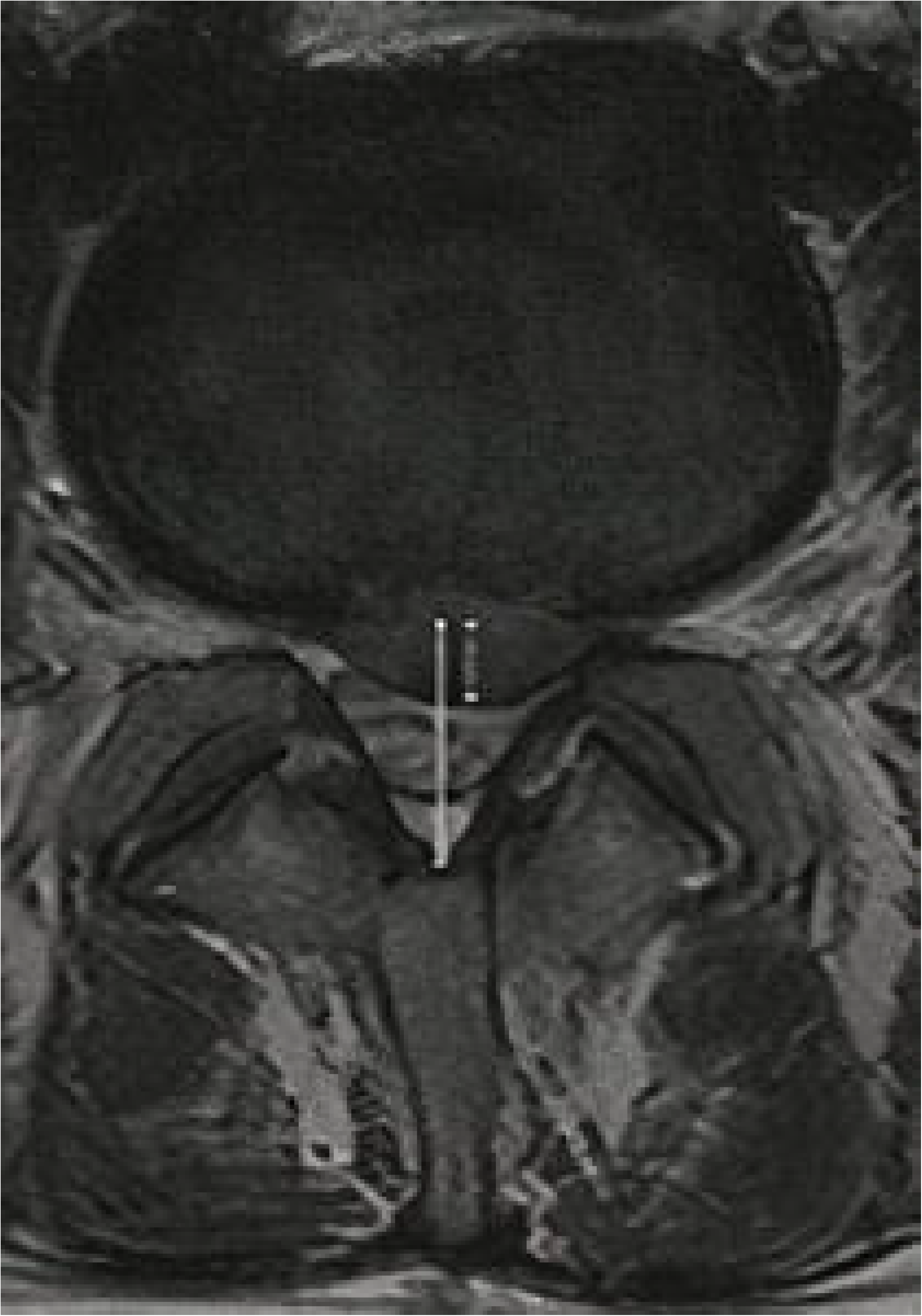
Measurements of the horizontal diameter of the disk herniation (dashed line) and the horizontal diameter of the spinal canal (continuous line) in the axial plane.

### Prednisone treatment protocol

Patients selected according to the above-mentioned criteria will undergo treatment with prednisone, a synthetic corticosteroid with little mineralocorticoid activity. It is the most widely available oral steroid,^36^ and the risk of suppression of the hypothalamic–adrenal axis is minimal with supraphysiological doses used for less than 3 weeks.^37^

Prednisone will be administered as 20mg tablets. This drug is available at low government-subsidized prices in many drugstores in the State of São Paulo, and patients will be advised to purchase the medication from participating drugstores, as is already routinely done regardless of participation in the research protocol. This means that the patients will not incur any additional costs for participating in this project.

The prescribed dosage of the medication will depend on the patient’s weight, since prednisone’s metabolism and distribution volume are related to body mass.

Therefore, patients who weigh more than 50 kg will receive the following administration regimen, similar to those routinely use and employed in previous studies:

1. 60 mg (3 tablets) in the first 5 days
2. 40 mg (2 tablets) in the next 5 days
3. 20 mg (1 tablet) in the following 5 days
4. 10 mg (1/2 tablet) in the last 5 days

The cumulative dose in this scheme is 650 mg, which is sufficient to promote an anti-inflammatory effect with minimal risk of immunosuppression.^37^

Patients under 50 kg will be prescribed the following dosage, in order to achieve the same objectives without risk of undesirable side effects:

1. 40 mg (2 tablets) for the first 10 days
2. 20 mg (1 tablet) for the next 5 days
3. 10 mg (1/2 tablet) in the following 5 days

### Concomitant use of other medication

Simple analgesic and muscle relaxant medications will be prescribed for immediate relief if the pain is very intense. NSAIDs use will be restricted due to its corticosteroid-like effect. These data will also be reported and considered for analysis by comparing the cumulative dose of those drugs between both groups.

### Notification of adverse effects

Each participant in this trial will receive guidance on possible adverse effects that may be related to treatment. This trial will use the definitions of adverse event (AE) and severe adverse event (SAE) established by the Clinical Safety Data Management guidelines.

The occurrence of any SAE will be reported to the study team through either a telephone channel made available to the patients or a face-to-face consultation at the SPH emergency room.

Minor AEs that do not meet the SAE definition will be reported during outpatient follow-up and annotated in the patient’s medical record and in the study form.

Those who doesn’t take medications as oriented will be classified as therapeutic failure and the reasons are going to be registered.

### Study schedule

#### Initial interview—first consultation

The data of potentially eligible patients in follow-up at the Orthopedic and Spine outpatient clinic will be annotated, and the main author of this study will be notified about it. Information such as telephone number, registration number, and home address will be recorded. Additionally, imaging studies such as anteroposterior and lateral radiography and lumbosacral MRI of the lumbar spine will be scheduled within one month after the first evaluation.

The patient will then be clinically evaluated by the authors on the same day of the first contact or during the following week, after orientation on participation in the study and signing of the ICF.

During the first consultation, demographic and clinical data will be obtained by one of the authors, as specified in previous sections. Additionally, the VAS and ODI questionnaires will be applied.

The treatment scheme with prednisone, as described in item treatment protocol section, will also be prescribed during the same consultation.

The patient will be scheduled for a return outpatient consultation after the end of the course of treatment with prednisone, i.e., after 20 days.

### Second consultation

The second consultation will occur after the course of treatment with prednisone, i.e, approximately 20 days, and will be conducted by the authors of this study.

During this consultation, the patient will fill the VAS and ODI questionnaires again, in order to quantify the improvement in relation to pain and function obtained with the proposed treatment.

Patients will be considered to have responded to treatment if they achieved a 50% reduction in VAS score or a 7.1% reduction in ODI score, according to parameters established in previous studies that considered these values as minimum clinical important difference (MCID).^32,33^

Thereafter, the parameters that had been obtained during the first consultation of those patients who responded to treatment will be compared to those of patients with no effective results, in order to identify statistically relevant variations of these parameters and, consequently, to identify possible predictors of success of treatment with prednisone.

Adherence to treatment among participants and any AEs they may have presented will also be assessed. Additionally, the resumption of normal activities and restrictions that still persist will be evaluated in relation to time.

### Third consultation

A third consultation will be performed 2 months (60 days) after the first consultation.

This consultation will be conducted by the study team, and the patient will again answer the ODI and low back pain/lower limb VAS questionnaires.

The main objective of this third consultation will be to assess whether the patients whose primary response was satisfactory maintained the improvement in function and pain and if a worsening of symptoms is reported, to identify which parameters may be related to good long-term response to the treatment.

### Statistical analysis and evaluation of results

The primary aim of the study is to compare social, clinical, and demographic parameters between the groups with favorable and unfavorable results, in relation to pain and function improvement, after the course of treatment with prednisone.

Initially, patients will be distributed into two groups: responsive and non-responsive, according to the improvements in pain and function obtained, as assessed by ODI and VAS scores. The mean value of the standard deviations of ODI scores described in the literature for similar studies was 15.1%.

Thereafter, both groups will be compared in relation to a total of 19 parameters: 6 demographic, 7 clinical, and 6 imaging parameters. Of these, 13 are categorical variables and 6 are continuous variables. A chi-square test will be applied for the categorical variables and a paired Wilcoxon test for continuous parameters.

Then, multivariable logistic regression methods will be applied to obtain the odds ratio with a confidence interval ≥ 95% for each parameter that presented statistically relevant differences between the groups, thereby assessing each parameter’s impact on effectiveness. These regression analyses aim to avoid biases of multivariable interaction.

A relatively low value of 10% loss to follow-up is estimated due to the fact that the primary outcome of the study will be obtained in a relatively short interval after primary care. Therefore, to obtain results that respect the ≥ 95% confidence interval, a sample of 300 patients is required. It is expected that 20–25 patients per month will be recruited from both outpatient clinics; therefore, the study is expected to be concluded in approximately 2 years, including the time required for the analysis of the primary outcome.

The cross-sectional epidemiological evaluation will be done through a contingency table, comparing the patients’ epidemiological data according to the degree of pain and loss of function. A chi-square test and Fisher’s exact test will be used for statistical analysis.

## Data Availability

The authors declare that the final dataset generated will be stored in a non-publically available repository (redcap.epm.br). All the sociodemographic, clinical and procedures will be available for researchers under request.

https://redcap.epm.br

## ETHICS AND DISSEMINATION

The study received the approval of Federal University of São Paulo Research Committee (4.232.193) and Research Ethics National Committee (CONEP).

The final dataset generated will be stored in a non-publically available repository (red-cap.epm.br). All the sociodemographic, clinical and procedures will be available for researchers under request. The final results will be submitted for publications in a peer-reviewed journal and conference presentation.

### Informed consent

In accordance with the rules of the Brazilian National Research Ethics Commission, an ICF will be provided to all patients recruited for the study. The ICF will be written and will contain the objectives and expected results of the present trial.

Additionally, the risks and benefits will be clarified. After completion of reading, the patients will be encouraged to ask questions and, if participation is amenable to the patients, they will be asked to sign the form. The patient will keep a copy of the ICF.

The Ethics Committee’s approval opinion and the Informed Consent Form that will be presented to the participants are attached to the present document.

## AUTHOR CONTRIBUTIONS

The study concept and design were conceived by GHP, RSU and MJT. The participants screening and data collection will be performed by GHP, CAC, FAV, AFF and DDC, EBP, RSU and MJT provided the feedback of the study design and data analysis. All authors provided edit and critical intellectual input to the manuscript, approving the final version.

